# Dispersal history of SARS-CoV-2 in Galicia, Spain

**DOI:** 10.1101/2024.02.27.24303385

**Authors:** Pilar Gallego-García, Nuria Estévez-Gómez, Loretta De Chiara, Pilar Alvariño, Pedro M. Juiz-González, Isabel Torres-Beceiro, Margarita Poza, Juan A. Vallejo, Soraya Rumbo-Feal, Kelly Conde-Pérez, Pablo Aja-Macaya, Susana Ladra, Antonio Moreno-Flores, María J. Gude-González, Amparo Coira, Antonio Aguilera, José J. Costa-Alcalde, Rocío Trastoy, Gema Barbeito-Castiñeiras, Daniel García-Souto, José M. C. Tubio, Matilde Trigo-Daporta, Pablo Camacho-Zamora, Juan García Costa, María González-Domínguez, Luis Canoura-Fernández, Daniel Glez-Peña, Sonia Pérez-Castro, Jorge J. Cabrera, Carlos Daviña-Núñez, Montserrat Godoy-Diz, Ana Belén Treinta-Álvarez, Maria Isabel Veiga, João Carlos Sousa, Nuno S. Osório, Iñaki Comas, Fernando González-Candelas, Samuel L. Hong, Nena Bollen, Simon Dellicour, Guy Baele, Marc A. Suchard, Philippe Lemey, Andrés Agulla, Germán Bou, Pilar Alonso-García, María Luisa Pérez-del-Molino, Marta García-Campello, Isabel Paz-Vidal, Benito Regueiro, David Posada

## Abstract

The dynamics of SARS-CoV-2 transmission are influenced by a variety of factors, including social restrictions and the emergence of distinct variants. In this study, we delve into the origins and dissemination of the Alpha, Delta, and Omicron variants of concern in Galicia, northwest Spain. For this, we leveraged genomic data collected by the EPICOVIGAL Consortium and from the GISAID database, along with mobility information from other Spanish regions and foreign countries. Our analysis indicates that initial introductions during the Alpha phase were predominantly from other Spanish regions and France. However, as the pandemic progressed, introductions from Portugal and the USA became increasingly significant. Notably, Galicia’s major coastal cities emerged as critical hubs for viral transmission, highlighting their role in sustaining and spreading the virus. This research emphasizes the critical role of regional connectivity in the spread of SARS-CoV-2 and offers essential insights for enhancing public health strategies and surveillance measures.

## Introduction

SARS-CoV-2 (severe acute respiratory syndrome coronavirus 2), a member of the betacoronavirus family, was first identified in December 2019 in Wuhan, China (Worobey et al. 2022). It is responsible for the coronavirus disease (COVID-19) pandemic, which was declared a public health emergency of international concern by the World Health Organization (WHO) on the 30th of January 2020 (Eurosurveillance editorial team 2020). As of June 2023, there have been 690 million people globally infected with SARS-CoV-2, resulting in more than 6.8 million confirmed fatalities. In response, nations worldwide enforced a variety of containment measures, ranging from social distancing and mandatory mask-wearing to lockdowns and, subsequently, vaccination campaigns (Sharma et al. 2021; Baniasad et al. 2021). In Spain, the virus affected over 13.8 million individuals and was linked to approximately 120,000 deaths (Worldometer 2020). While some containment strategies in Spain, such as national lockdowns and mask mandates, were uniform across the country, others were implemented at the discretion of the various autonomous regions or territories (“Plan de Respuesta Temprana En Un Escenario de Control de La Pandemia Por COVID-19” 2020).

In this study, we explore the historical dispersal of SARS-CoV-2 within Galicia, an autonomous community located in the northwest corner of the Iberian Peninsula, bordering Portugal. Endowed with independent governance, particularly in health matters, Galicia segments its healthcare services across seven main districts, each coupled with a major public hospital servicing the seven principal cities (Supplementary Figure 1). A Coruña and Vigo are the most populous, while Santiago de Compostela is the regional capital and a renowned pilgrimage destination. Besides these urban centers, smaller towns and rural areas account for 63.3% of the region’s population, highlighting its disseminated population structure (Galician Institute of Statistics, https://www.ige.gal).

By March 2022, Galicia, reflecting the wider scenario across Spain, had experienced six distinct waves of COVID-19 since the beginning of the pandemic (Figure 1). The initial wave, commencing around March 2020, was not distinguished by the number of cases but rather by the severity of counteractions, notably a nationwide lockdown. This phase gave way to a second wave in the summer of 2020, driven by relaxed restrictions and the spread of the B.1.177 lineage across Europe (Hodcroft et al. 2021). The Alpha variant of concern (VOC) entered Galicia and Spain during a third wave, proliferating alongside pre-existing variants. This surge was presumably fueled by increased social interactions over the Christmas period, despite the implementation of several containment strategies.

**Figure 1.**
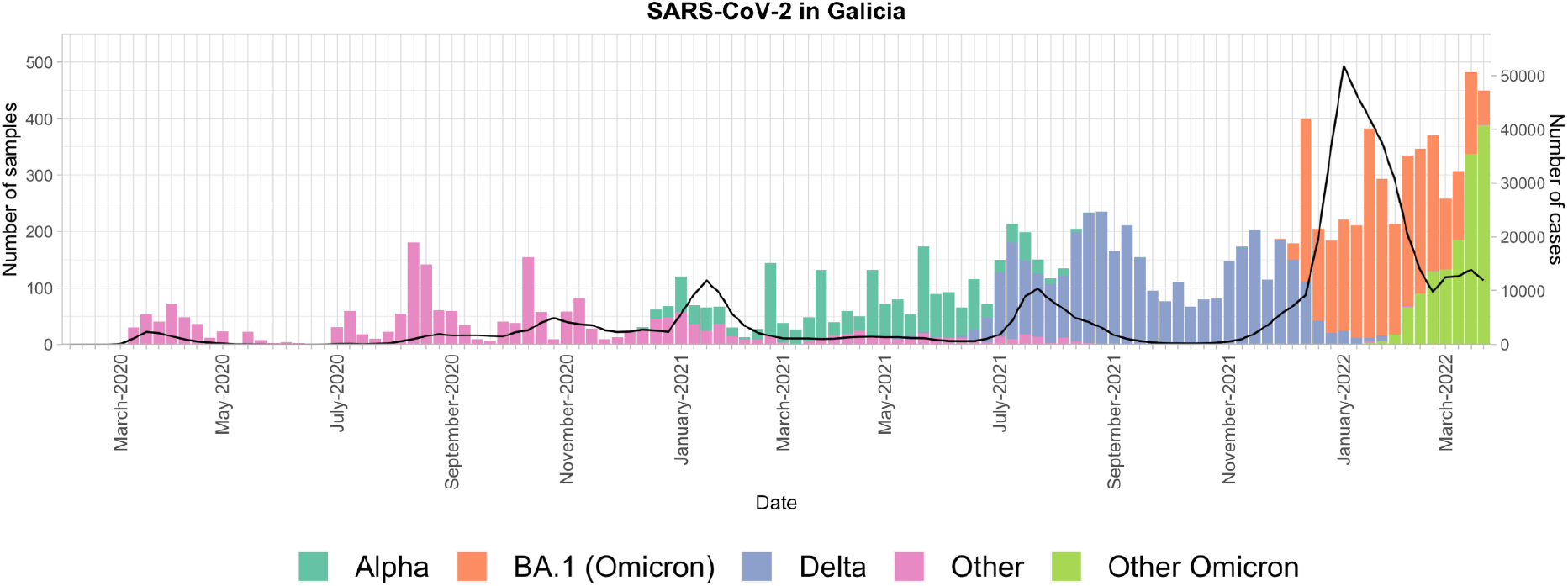
SARS-CoV-2 epidemic waves in Galicia from February 2020 to March 2022. Number of samples available in the GISAID database (left-axis; bars) and diagnosed cases recorded by the Spanish National Epidemiology Center (CNE) (right-axis; black line) for Galicia from February 2020 to March 2022. Bar colors represent different lineages or variants, as explained in the legend.

The fourth wave was primarily dominated by the Alpha variant but also saw non-pervasive introductions of other VOCs, such as Beta and Gamma. This period coincided with the initiation of the vaccination campaign and the reinstatement of rigid public health guidelines, such as the virtual delivery of university courses in Galicia, resulting in fewer cases. The emergence of the Delta variant in the summer of 2021 propelled Galicia into its fifth epidemic wave, marked by a significant surge in cases. Finally, the sixth wave, beginning in October 2021 and pushing into early 2022, encompassed the ending of Delta and the ascent of Omicron, facilitating the highest number of cases across the region during the pandemic. This occurred amidst the least stringent restrictions to date, with a short-lived reinstatement of mandatory outdoor mask-wearing.

To delve into the progression of the SARS-CoV-2 epidemic within Galicia, we formed the EPICOVIGAL Consortium (https://epicovigal.uvigo.es). This consortium represents a collaborative endeavor comprising a network of laboratories affiliated with three universities and the Microbiology Departments from the seven primary hospitals under the Galician Public Health System (Servizo Galego de Saúde, SERGAS). This strategic partnership was pivotal from March 2020 through June 2022, during which the consortium successfully sequenced 4,199 SARS-CoV-2 samples. Out of these, 2,471 samples met our stringent quality criteria and were identifiable as belonging to what we have designated as the study’s focus: the Alpha, Delta, and Omicron-BA.1 variants. Our investigational approach hinged on a Bayesian phylogeographic paradigm, selected for its robustness in handling complex data sets. This methodology enabled us to examine 1,182 sequences derived from our consortium’s efforts, in conjunction with those sourced from global and Spanish sequences available in public databases.

## Methods

This study was conducted by members of the EPICOVIGAL Consortium under the approval of the Research Ethics Committee of Pontevedra-Vigo-Ourense (code 2020/422).

### Sample collection

Nasopharyngeal exudate samples from individuals putatively infected with SARS-CoV-2 were selected by the EPICOVIGAL members of the Microbiology Departments of the following hospitals of the Public Health System in Galicia (Servizo Galego de Saúde, SERGAS): Complexo Hospitalario Universitario de Ferrol (CHUF), Complexo Hospitalario Universitario de A Coruña (CHUAC), Hospital Universitario Lucus Augusti (HULA), Complexo Hospitalario Universitario de Santiago de Compostela (CHUS), Complexo Hospitalario Universitario de Ourense (CHUO), Complexo Hospitalario de Pontevedra (CHOP), and Complexo Hospitalario Universitario de Vigo (CHUVI). A total of 4,199 leftovers of the samples received for the routine diagnosis of SARS-CoV-2 infection were studied. We also collected metadata such as gender, age, town of residence, and postal code when possible. A detailed description of the samples obtained by the EPICOVIGAL Consortium can be accessed from the Nextstrain (https://nextstrain.org/groups/epicovigal/ncov/galicia-in-progress) and Microreact sites (https://microreact.org/project/nzya2sYASCUBxEkF24PV49-epicovigal) (Hadfield et al. 2018; Argimón et al. 2016).

### RNA extraction and viral load measurement

Following the manufacturer’s recommendations, we extracted DNA/RNA from the nasopharyngeal exudates with different kits (Supplementary Table 1). We measured each sample’s SARS-CoV-2 genome copy number by real-time RT-PCR of the ORF1ab, E, N, RdRP, or S genes (Supplementary Table 1). Only samples with Ct < 30 were analyzed further.

### cDNA synthesis and multiplex amplification

We followed the ARTIC sequencing protocol (v.3) (Quick et al. 2017) with slight modifications. This multiplex PCR-based target enrichment produces 400 bp amplicons that span the SARS-CoV-2 genome. First, we retrotranscribed the RNA samples to cDNA using the SuperScript IV reverse transcriptase (Invitrogen, MA, USA), starting with 11 μL of RNA. Then, we ran 30-35 PCR cycles for all the samples, independently of the Ct value, using the ARTIC nCoV-2019 V3 (IDT, CA, USA) panel until October 2021 and the ARTIC V4 nCoV-2019 (IDT, CA, USA) panel afterward, with the Q5 Hot Start DNA polymerase (New England Biolabs, MA, USA). PCR products were mixed, cleaned, and eluted with 35 μL NFW (or 10 mM Tris-Cl, pH 8.5) and quantified with the Qubit 3.0 using the dsDNA BR or HS kits (Thermo Fisher Scientific, MA, USA). We checked the amplicon sizes with the 2200/4150 TapeStation D1000 kit (Agilent Technologies, CA, USA) or the 2100 Bioanalyzer System (Agilent Technologies, CA, USA).

### Library construction and genome sequencing

We built a single whole-genome sequencing library for each sample, employing the DNA Prep (M) Tagmentation kit (Illumina, CA, USA) using 1/4 of the recommended volume, with approximately 100-125 ng of input DNA. We quantified the sequencing libraries with the Qubit 3.0 using the dsDNA HS kit (Thermo Fisher Scientific, MA, USA) and checked their sizes with the 2200/4150 TapeStation D1000 kit (Agilent Technologies, CA, USA) or with the 2100 Bioanalyzer System (Agilent Technologies, CA, USA). Finally, we sequenced them on the iSeq (PE150 reads), MiniSeq (PE150 reads), and Miseq (PE300 reads) platforms at the sequencing facilities of CHUAC, CHUS, CHUVI and University of Vigo.

### Construction of consensus sequences

For each of the resulting FASTQ files, we used FastQC v.0.11.9 (Andrews 2010) to assess the quality of the reads. We mapped the reads to the reference SARS-CoV-2 genome MN908947.3 using BWA v.0.7.17 (Li 2013) and trimmed them using iVar v.1.3.1 (Grubaugh et al. 2019) (minimum length of the read after trimming = 30; minimum quality threshold for the sliding window = 20; width of the sliding window = 4). After trimming, we used iVar to generate the consensus sequence for each sample (minimum depth = 10; minimum frequency threshold = 0.5; minimum quality score threshold per base = 20). We assessed the quality of the consensus sequences and classified those with “good” overall status into clades with Nextclade (v.2.3.1) (Aksamentov et al. 2021) and into PANGO lineages with Pangolin v.4.1.3 (O’Toole et al. 2021). The final number of Alpha, Delta, and Omicron (BA.1) consensus sequences that passed all the filters was 2,471.

### Collection of additional Galician sequences from GISAID

We downloaded all the Galician sequences available in GISAID (Elbe and Buckland-Merrett 2017) for the Alpha, Delta, and Omicron VOCs that met the following GISAID criteria: were “complete” (length > 29,000 nt); presented a complete collection date (year-month-day); came from a human; and, for Alpha and Delta, were “high coverage” (<1% of missing bases and <0.05% of unique amino acid mutations). For Omicron, we excluded those with “low coverage” (>5% missing bases) but did not enforce the “high coverage” filter, as it would imply discarding >90% of the Omicron samples from all around the world. For each of these three VOCs, we retained only those samples with “good quality” according to Nextclade and that were successfully assigned to the corresponding Nextclade clade or PANGO lineage (clade 20I / lineage B.1.1.7 for Alpha and clades 21A, 21I & 21J / lineage B.1.617.2 and sublineages (AY) for Delta). For Omicron, we only considered lineage BA.1 and its sublineages (clade 21K), as the pandemic wave corresponding to BA.2 and the rest of the Omicron sublineages was still ongoing at the start of this study. The final number of Alpha, Delta, and Omicron (BA.1) Galician sequences in GISAID that passed all the filters was 6,964.

### Selection of non-Galician sequences

To estimate the history of the introductions of Alpha, Delta, and Omicron-BA.1 in Galicia from other Spanish regions or countries, we sampled several non-Galician SARS-CoV-2 sequences from GISAID according to their prior probability of resulting in novel introductions in Galicia. We used mobility data from the Spanish National Institute of Statistics (INE; https://www.ine.es) to calculate the latter, corresponding to the number of mobile phones entering different Spanish regions and the region or country of origin. We note that any type of mobility data presents limitations, depending on how it is obtained, and it may not be representative of the whole population, as not everyone carries a mobile phone, for example. However, it still provides a good approximation of the influx of travelers in and out of specific locations (Lemey et al. 2021; du Plessis et al. 2021). We considered each Spanish region as different entities at the same level as countries. We sampled sequences between weeks 52 of 2020 and 32 of 2021 (2020-12-20 to 2021-08-14) for Alpha, between weeks 25 and 50 of 2021 (2021-06-20 to 2021-12-18) for Delta, and between weeks 49 of 2021 and 13 of 2022 (2021-12-05 - 2022-04-02) for BA.1. We computed the connectivity from all the available locations to Galicia for each of these periods, ranked them and selected the regions and countries that represented the 95% of the mobility to Galicia. We computed the number of samples to collect from each location as the *number of reported cases in the region/country* × *fraction of connectivity to Galicia* × *6*.*5/10,000*, with a minimum number of samples of 100 for Alpha and Delta and 50 for Omicron-BA.1. These thresholds were arbitrarily set to obtain large but manageable data sets, as in (Truong Nguyen et al. 2022). When possible, we selected an even number of samples per week and distinct locations for each region or country. Alpha and Omicron-BA.1 sequences were downloaded on 2022-06-11, while the Delta sequences were downloaded on 2022-04-26. We retained sequences with a “good” overall status in Nextclade and removed duplicates with seqkit v2.1.0 (Shen et al. 2016).

With these sequences, we built a multiple sequence alignment with Nextalign v.1.11.0 (Hadfield et al. 2018) for each VOC, including a random sample of 1,000 Galician sequences. For each alignment, we estimated a maximum likelihood (ML) tree with IQ-TREE v.2.1.3 (Minh et al. 2020) based on the optimal substitution model suggested by ModelFinder (Kalyaanamoorthy et al. 2017). Nodal support was computed with 2,000 ultrafast bootstrap replicates (Hoang et al. 2018). We used TreeTime v.0.8.1 (Sagulenko, Puller, and Neher 2018) with the option “clock” to detect temporal outliers and removed them from further analyses. The final number of sequences per alignment was 3,695, 3,548, and 3,529 for Alpha, Delta, and Omicron-BA.1.

### Bayesian analysis of introductions

We used BEAST v1.10.5 (prerelease #23570d1) (Suchard et al. 2018) to infer the VOC introductions. BEAST employs a formal Bayesian framework that considers the uncertainty of the phylogeny while estimating several phylodynamic parameters of interest. Here, we leveraged a phylogeographic generalized linear model (GLM) incorporating the region/country of sampling as a discrete trait and including a connectivity matrix between locations as a covariate (Gill et al. 2016; Lemey et al. 2021). This matrix was log-transformed and standardized before being included in the phylogeographic GLM. We used the INE mobility data previously described for the connectivity data between Spanish regions and between them and other countries. For the connectivity between countries, we used aggregated Facebook international mobility data from January 2021 to March 2022. We opted to use Facebook data instead of Google aggregated mobility data because it was more readily available for our period of interest. Facebook mobility data correlates highly to Google mobility data (Lemey et al. 2021).

We started our Markov chain Monte Carlo (MCMC) analysis in BEAST v1.10.5 using a timed tree estimated by TreeTime (with options --max-iter 10 and --reroot least-squares) as starting tree, using HKY as the substitution model (Hasegawa, Kishino, and Yano 1985), with empirical base frequencies, and rate variation among sites following a discretized gamma distribution with four rate categories (Yang 1994), with a fixed evolutionary rate of 7.5 × 10^-4^ nucleotide substitutions/site/year (du Plessis et al. 2021) and a Bayesian non-parametric coalescent (skygrid) model (Gill et al. 2013) as the tree prior. Missing covariate values in the connectivity matrices were integrated out using efficient gradient approximations (Didier et al. 2024) to the trait data likelihood (Ji et al. 2020). We simulated seven or more independent MCMC chains per dataset, discarded at least 10 percent of each chain as burn-in, and combined the results with LogCombiner (Suchard et al. 2018). We ran the chains until all continuous parameters (or most in the case of Omicron-BA.1, see below) presented effective sampling sizes (ESSs) > 100, totaling 1,214M generations for Alpha and 1,500M for Delta. For the Omicron-BA.1 analysis, which has a higher location state dimensionality and hence a larger number of missing covariate values, 3.125% of missing covariate values presented an ESS < 100 after 1,200M combined generations. From the posterior tree distributions, we summarized the transition histories and their timing using the tool TreeMarkovJumpHistoryAnalyzer implemented in BEAST (Minin and Suchard 2008; Lemey et al. 2021). All the BEAST analyses were executed using the BEAGLE library (v.4) (Ayres et al. 2019).

### Identification of Galician transmission clusters

We used a three-step approach to identify specific outbreaks and transmission clusters in Galicia. First, we used Breakfast (Huska and Beslic 2022) to delimit clusters of closely related sequences. Breakfast is a simple method for clustering SARS-CoV-2 genomes using sets of mutations (e.g., detected by Nextclade) that can work with large datasets. Second, we reconstructed the phylogenetic relationships within these clusters using all the Galician sequences and a random sample of non-Galician sequences. Third, we estimated the most likely location of all the ancestral nodes and identified monophyletic groups of Galician sequences sharing a most recent common ancestor in Galicia (see below). For this purpose, we downloaded all the available sequences from GISAID for Alpha and Omicron on 2022-06-11 and Delta on 2022-05-13. After removing duplicates and low-quality sequences, we added to these datasets all the available Galician sequences in GISAID to those we sequenced. We ran Breakfast v.0.4.3 with a minimum size of 5 to call a cluster and a maximum pairwise distance of 1 between clustered sequences without skipping deletions or insertions. In the three epidemic waves under study, there was a single Breakfast cluster that included most of the sequences (51% of Alpha, 91% of Delta, and 99% of Omicron-BA.1). These three clusters contain millions of sequences, so we did a random subsampling of the non-Galician sequences in these large clusters, keeping an even number of sequences per week. Including the Galician sequences, the final datasets contained ∼2,000 sequences for Alpha (the smallest cluster) and ∼10,000 for Delta and Omicron. We estimated maximum likelihood (ML) trees for these three datasets using IQ-TREE under the optimal substitution model suggested by ModelFinder, with 1,000 ultrafast bootstrap replicates. Then, we carried out an ML ancestral reconstruction of the character location (“Galicia” or “Other”) at the internal nodes of these trees with the “ape” package (Paradis and Schliep 2019) in R (R Core Team 2022). Finally, we extracted the clusters in those trees exclusively composed of Galician sequences that shared a parental node assigned to “Galicia”. We repeated the random subsampling described above and the whole cluster identification procedure five times to ensure that the Galician clusters identified were independent of the subsampling (Kaleta et al. 2022). Apart from these three large clusters, there were smaller clusters containing only Galician samples that we used in full and others that were not informative enough to analyze (i.e., clusters in which all the samples belonged to the same location and lacked zip codes).

### Bayesian phylogeographic reconstruction

We aligned the sequences using Nextalign for each transmission cluster and inferred the ML tree using IQ-TREE. We used BEAST to infer the phylogeographic history of each transmission cluster. For this, we included the geographical coordinates for each sequence as a trait. These coordinates were extracted from the sample’s location or zip code. To avoid two samples being associated with the same coordinates, we added a jitter (0.001) to each value (Dellicour et al. 2022, 2016). We started the MCMC analysis on the ML tree estimated by IQ-TREE, using HKY as the substitution model, with empirical base frequencies and rate variation among sites following a discretized gamma distribution with four rate categories, with a fixed evolutionary rate of 7.5 × 10^-4^ and a Bayesian non-parametric coalescent model as the tree prior. We simulated multiple independent MCMC chains per dataset and combined the results with LogCombiner. We ran the MCMC analyses until all parameters presented combined ESSs > 200 and built a maximum clade credibility (MCC) tree using TreeAnnotator v.1.10.4 (Suchard et al. 2018) with 1,000 samples from the posterior distribution. Finally, we used the R package “seraphim” (Dellicour et al. 2016) to extract the spatiotemporal information embedded in the MCC tree and to generate a graph of the dispersal history for each cluster.

## Results

### Introductions of SARS-CoV-2 VOCs in Galicia

Our analysis revealed at least 96 independent introductions (95% highest posterior density [HPD] = 84-106) of the Alpha variant in Galicia. Among these, 83 (95% HPD = 73-92) had a “low” (≤10) number of sampled descendants, including 40 (95% HPD = 33-46) with only one descendant (i.e., singletons). Most of these introductions were inferred to originate from other Spanish regions (Figure 2A-B), particularly Madrid, Castile and Leon, Catalonia, and Asturias. Interestingly, the number of introductions from France ranked second, only after those from Madrid. Additionally, we identified five introductions from Portugal, a neighboring country that shares an extensive border with Galicia. Thirteen international introductions resulted in a noticeable number (>10) of Galician descendants, including one introduction from France around November 2020, which produced 194 descendants. This event, occurring just before the surge of the Alpha wave within Galicia, might have had a relevant influence on the proliferation of this variant across the region.

**Figure 2.**
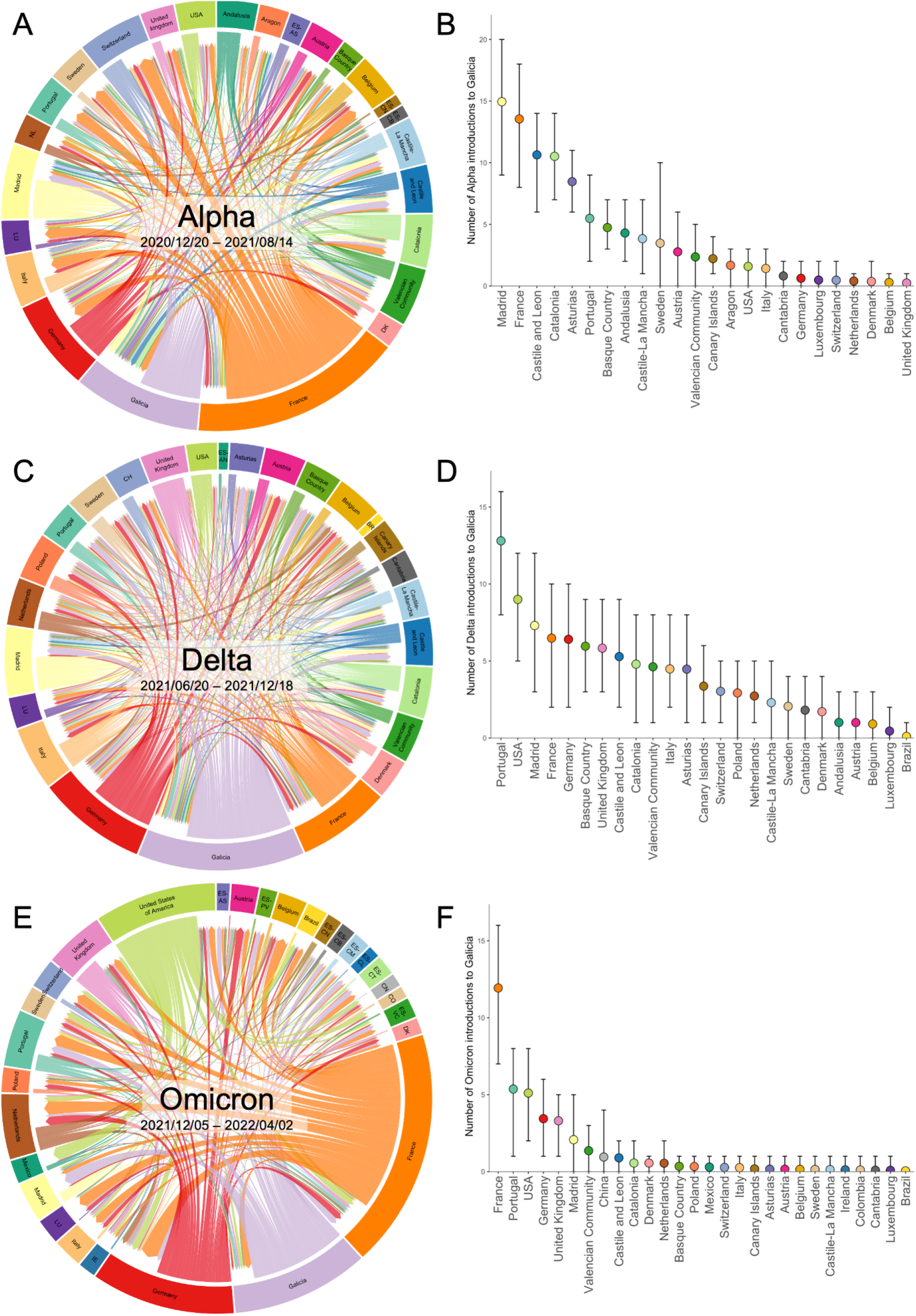
SARS-CoV-2 introductions into Galicia during the Alpha wave. Left: Circular migration flow plot based on the Markov jumps between multiple countries and several Spanish regions, including Galicia. (A: Alpha; C: Delta; E: Omicron). Right: Mean and 95% highest posterior density (HPD) number of transitions from Spain and other countries to Galicia, computed from all the trees from the posterior distribution. (B: Alpha; D: Delta; F: Omicron). Correspondence between codes ISO 3166-2 and the complete name of the location in Supplementary Table 2.

For the Delta variant, we inferred 101 (95% HPD = 87-112) independent introductions (Figure 2C-D). Like Alpha, most Delta introductions were also small, with only 11 resulting in more than 10 sampled descendants. However, the origins of the Delta introductions differ from those observed with the Alpha variant, with Portugal and USA being the most significant sources, followed by Madrid, France, and Germany. The emergence of the Delta wave in Galicia occurred during the summer of 2021, coinciding with the relaxation of previous containment measures and an increase in international tourism, which likely facilitated the spread of this variant. Specifically noteworthy were two Delta introductions from the United Kingdom (UK) –a first one in early June 2021 and a subsequent one in mid-July 2021– leading to the identification of 31 (95% HPD = 23-36) and 67 (95% HPD = 66-67) sampled descendants, respectively. During the initial wave of the Omicron variant (encompassing BA.1 and its sublineages) in Galicia, we identified 39 introductions (95% highest posterior density [HPD] = 28-46) (Figure 2E-F), notably lower than the number of introductions attributed to Alpha and Delta. Consistent with the patterns observed for Alpha and Delta, most Omicron introductions left a limited number of descendants, with only four (95% HPD = 2-6) leaving more than 10. Intriguingly, despite not overlapping with the summer vacation period –a peak time for international travel–, most Omicron introductions in Galicia were predominantly international, particularly from France, Portugal, the USA, Germany, and the UK.

### Phylogeography reconstructions

We identified five transmission clusters that suggested significant local spread: five for Alpha, two for Delta, and a single cluster for Omicron. These clusters were selected for more in-depth exploration based on two criteria: each contained at least twenty Galician samples and showcased a geographical distribution, spreading across different locations or associated with distinct zip codes.

Most of the selected Alpha clusters showed relatively high mobility and connectivity across Galicia. Cluster 1-1 (Figure 3A) has an uncertain origin, with ancestors in the three main Galician cities, Vigo, Santiago de Compostela, and A Coruña. These results suggest sustained migration over time not only within these cities but also among them. Cluster 1-2 (Figure 3B) and Cluster 2 (Figure 3C) appear to have originated in Vigo, in the southwest, and later spread to more remote areas. Cluster 3 (Figure 3D) likely originated in Santiago de Compostela, extending primarily to smaller adjacent locations but also more distant ones near A Coruña and the western part of Galicia. On the other hand, Cluster 4 (Figure 3E) is centered around A Coruña, with relatively few connections with neighboring locations.

**Figure 3.**
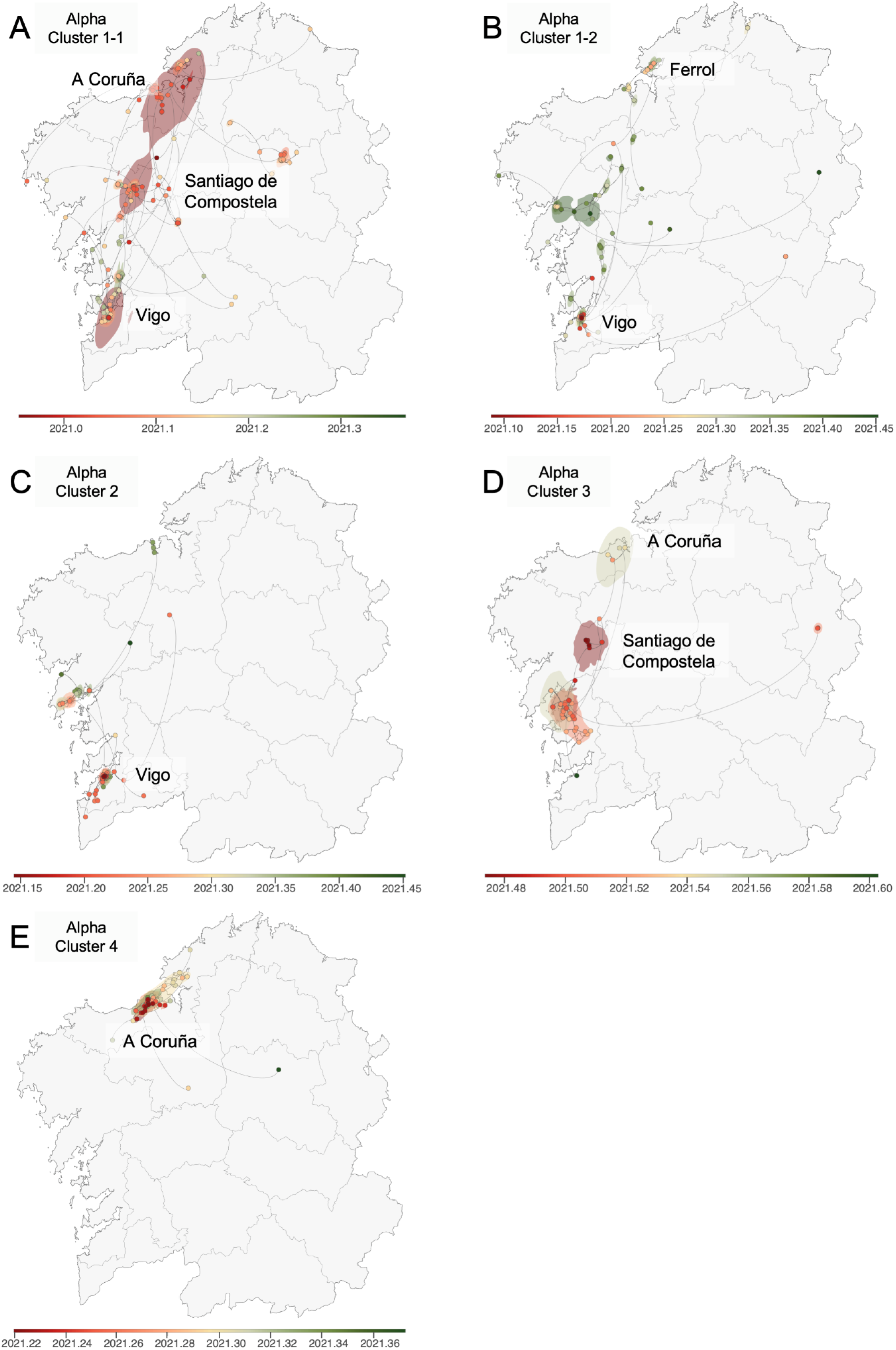
Continuous phylogeographic reconstruction of selected Alpha clusters. A) Cluster 1-1; B) Cluster 1-2; C) Cluster 2; D) Cluster 3; E) Cluster 4. We map the maximum clade credibility (MCC) trees and overall 80% highest posterior density (HPD) regions reflecting the uncertainty related to the Bayesian phylogeographic inference. MCC trees and 80% HPD regions are based on 1,000 trees sampled from each posterior distribution and colored based on their time of occurrence. The direction of viral lineage movement is indicated by edge curvature (anti-clockwise).

In the case of Delta, cluster 1-1 (Figure 4A and 4B) consists of sequences almost exclusively from Vigo. The lack of a discernible clustering pattern within the city itself is interesting. Still, this ambiguity regarding the virus’s spread within Vigo could stem from limitations in pinpointing exact locations due to the use of postal codes and jittered coordinates). An additional factor could be the lifting of several COVID-19 restrictions, including those impacting nightlife venues. In contrast, subcluster 1-2 (Figure 4C) exhibits a significantly broader dispersion across the entirety of Galicia, again marking the region’s three largest cities as primary hotspots, with a likely origin in Vigo.

**Figure 4.**
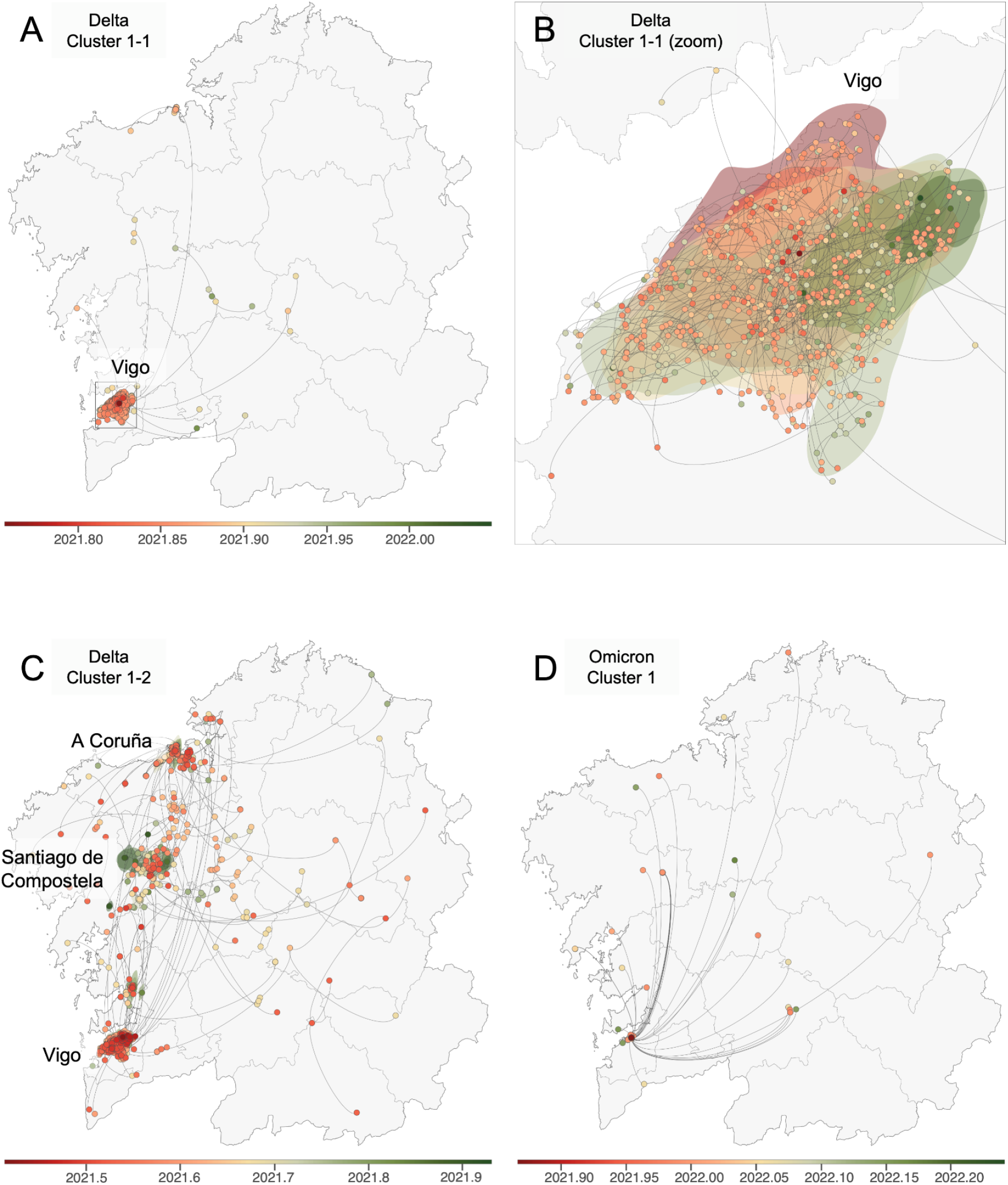
Continuous phylogeographic reconstruction of selected Delta clusters and Omicron - BA.1 cluster 1. A) Delta Cluster 1-1, general vision; B) Delta Cluster 1-1, zoom in Vigo; C) Delta Cluster 1-2; D) Omicron - BA.1 Cluster 1. We map the maximum clade credibility (MCC) trees and overall 80% highest posterior density (HPD) regions, reflecting the uncertainty related to the Bayesian phylogeographic inference. MCC trees and 80% HPD regions are based on 1,000 trees sampled from each posterior distribution and colored based on their time of occurrence. The direction of viral lineage movement is indicated by edge curvature (anti-clockwise).

The Omicron cluster (Figure 4D) arose in Vigo and mainly circulated inside the city, with some connections to other locations, but apparently without further local transmission.

## Discussion

The dynamics surrounding the introductions of the SARS-CoV-2 variants into Galicia underscores the complexities tied to geographical, social, and structural factors within regions. Galicia’s position in the northwest corner of Spain, bordered by Portugal to the south and the Atlantic Ocean to the west and north, fundamentally shapes its connectivity with both national and international regions. This peripheral location means terrestrial access is feasible only through other Spanish regions and Portugal, with air connections being relatively limited, especially during off-peak travel periods. These geographical conditions, together with the different non-pharmaceutical interventions implemented, which included perimeter closures during specific periods (i.e., the Holy Week), likely played a significant role in the pattern of SARS-CoV-2 introductions observed.

Most introductions of the Alpha variant into Galicia originated from other Spanish regions. Madrid accounted for the highest number of detected introductions during this period, likely due to its extensive connectivity with Galicia via numerous trains and flights. Given its status as Spain’s capital and Galicia as a favored holiday spot, considerable travel occurs between these regions for both business and leisure. This is evidenced by the detection of Galicia’s first COVID-19 case in 2020, originating from Madrid (Carballada and Balsa-Barreiro 2021). A similar scenario occurs with Catalonia, which, as Spain’s second-largest region in terms of population, maintains a strong connection to Galicia, facilitating the movement of individuals. The adjacency of Castile and Leon, and Asturias to Galicia further contributes to a notable number of introductions, underscored by their geographical proximity.

Most international introductions of the Alpha variant into Galicia were attributed to France, a country that, despite its relatively large distance from Galicia, maintains strong connections with Spain. Conversely, despite its geographical proximity and high connectivity with Galicia —including a significant daily cross-border workforce— Portugal accounted for a minimal number of Alpha introductions. This phenomenon is likely tied to the Spanish-Portuguese border closure from January 31 to May 1, 2021, allowing only crossings for justified reasons, such as work, whereas the Franco-Spanish border remained accessible during the same period. Introductions from other European countries may be accounted for by tourists or expatriates returning for the summer or holiday seasons. Interestingly, no introductions were detected from the UK, Alpha’s place of origin, which can likely be attributed to stringent travel restrictions imposed between the UK and Spain during this period (Kraemer et al. 2021). This observation underscores the influence of both travel policies and geographical connectivity on the pattern of SARS-CoV-2 variant introductions.

The observed increase in introductions during the Delta wave may be attributed to its onset in summer, a period marked by peak connectivity (Supplementary Figure 2), alongside a relaxation in several containment measures and Delta’s higher transmissibility compared to Alpha (Campbell et al. 2021). This phase witnessed the relaxation of stringent restrictions, including the lifting of perimeter closures and of the outdoor mask mandate. Concurrently, the acceleration of vaccination campaigns permitted the reopening of various leisure sectors, such as hostelry and nightlife, likely fueling international tourism. Notably, Portugal emerged as the principal origin of Delta introductions, closely followed by the USA. This contrasted with the Alpha wave period, characterized by more restrictive travel measures, limiting introductions largely to fewer countries. During the summer season, there is an increase in direct flights connecting Galician airports with several European destinations, which likely facilitated the observed increase in introductions. Additionally, domestic connectivity to Galicia peaked in August, mirroring the pattern of increased international connectivity. However, the number of national introductions during the Delta wave was comparatively smaller than in the Alpha period, underscoring the significant role of international movement in the spread of the Delta variant.

The onset of the Omicron-BA.1 wave was marked by the reinstatement of more stringent measures, coinciding with the Christmas holiday season. The enforcement of a COVID-19 certificate for access to various activities, recommendations for minimizing contacts, and the cancellation of New Year’s celebrations exemplify these tightened restrictions. By week 6 of 2022 (around February 10, 2022), outdoor mask mandates were lifted, and the requirement for the COVID-19 certificate was discontinued, aligning with a decrease in cases following the Christmas peak. The pattern of introductions during the Omicron wave diverged significantly from previous waves, with a total of 39 introductions, markedly less than those reported for Alpha and Delta (96 and 101, respectively). The introductions during this period were predominantly from international sources, with a smaller proportion of national ones. Despite increased connectivity associated with the Christmas season, it did not reach summer levels. However, the reduction in introductions during Omicron’s emergence cannot solely be attributed to the seasonal variation in connectivity, especially considering that total COVID-19 cases peaked during this period. This indicates that other factors, possibly including the implemented health measures and the characteristics of the Omicron variant itself, played significant roles in shaping the dynamics of its spread.

The three variants had in common that most introductions led to a low number of descendants, including a significant proportion of singletons, while only a minority resulted in extensive transmission chains. This pattern of transmission, characterized by the heterogeneity in the efficiency of SARS-CoV-2 to propagate and establish substantial local transmission networks, has also been described in other countries (Dellicour et al. 2021; Castelán-Sánchez et al. 2022).

The analysis of the Alpha clusters in Galicia revealed that control measures effectively reduced the virus’s spread from primary sources, predominantly situated in the largest cities. These containment strategies included the enforcement of municipal perimeter closures, restrictions on social gatherings, regulated operational hours or complete shutdown of hostelry, reduced capacities in retail establishments, and the transition to online teaching. Coupled with the inherently lower transmissibility of Alpha (Campbell et al. 2021), these measures likely played a crucial role in restricting the virus’s dissemination to other areas and minimized the extent of Alpha transmission clusters compared to those observed for Delta. The control measures were tightened immediately following the Christmas season, with a gradual relaxation as the summer approached. This phase saw the lifting of outdoor mask mandates and the reinstatement of nightlife activities, which likely contributed to heightened virus transmission to less populous areas towards the latter part of the analyzed period.

For the Delta wave, we detected a major transmission cluster that originated in some of the largest cities and subsequently dispersed to numerous remote areas. This development aligns with the timing of the Delta wave’s emergence at the tail end of the Alpha wave during the summer, a period characterized by the relaxation of various control measures. Conversely, the other Delta cluster exhibited a more localized behavior, primarily confined to the city of Vigo. This pattern reflects the relaxed containment strategies within urban centers, where the imposition of the COVID-19 certificate was mainly limited to regulating access to the interiors of venues.

The detailed analysis of the Omicron-BA.1 wave, particularly focusing on a transmission cluster centered in Vigo, revealed a pattern primarily consisting of within-city transmission alongside several long-distance transmission events that did not lead to further local outbreaks. During this period, traveling was discouraged, yet without an outright ban, and the implementation of various restrictions over the Christmas season potentially mitigated broader viral spread. The observed dynamics of this cluster, along with a relatively low number of detected introductions alongside Omicron BA.1’s higher transmissibility compared to Alpha and Delta (Elliott et al. 2022), and the peak in case numbers, presents a paradox. A potential explanation for this might lie in the limitations of sequencing capabilities. With an overwhelming number of cases, it is plausible that sequencing efforts could only target a selective number of cases from each outbreak, markedly less than in previous waves where a larger proportion of cases underwent sequencing due to lower overall numbers (Supplementary Figure 3). Consequently, this limitation may have impaired the detection of some introductions and clusters. Furthermore, Omicron BA.1’s tendency to result in a higher incidence of asymptomatic and mild cases, as influenced by preexisting immunity from vaccinations and past infections, and its own intrinsic properties (Carabelli et al. 2023; Bager et al. 2022), complicates direct comparisons with Alpha and Delta.

Throughout the emergence and spread of the three variants, connections between the main cities remained stable despite varying degrees of restrictions. This sustained interaction can be attributed to the fact that essential movements between cities, such as working, studying, or access to health services, were not restricted. Viral transmission within cities appeared steady and, despite intermittent stringent control measures, such as bans on meeting individuals from outside one’s household during the Alpha wave, essential activities at workplaces and educational institutions continued as potential venues for interactions. While masks were mandatory in Spain for long periods, they were not always correctly worn or used, contributing to ongoing transmission risks (Beca-Martínez et al. 2022). The majority of the identified transmission clusters were predominantly concentrated in coastal cities (A Coruña and Vigo) and the administrative capital (Santiago de Compostela), with much lower dissemination to inner locations. These cities also house the hospitals that contribute most Galician samples to the GISAID database (Supplementary Figure 4), suggesting a potential bias towards detecting clusters more frequently within these urban centers than in more remote or less populous areas.

Indeed, when interpreting these results, we must always consider the uneven sampling of the different Galician locations, the differential sequencing efforts, and the reduced sample sizes necessary to make different analyses computationally feasible, as all these elements can influence the results to some extent (Tegally et al. 2023; Tsui et al. 2023). Several strategies were employed in this study to mitigate the impact of these variables. Incorporating known connectivity data among different locations helped to contextualize the spread and potential transmission networks, providing a more informed framework for the analysis. Additionally, conducting multiple iterations of the analyses with subsamples helped to account for the variability introduced by the sample selection process. Despite these efforts, limitations stemming from the quantity and quality of the available sequence data and its metadata could still introduce biases, particularly for Omicron, where the fraction of cases that underwent sequencing was substantially lower.

In conclusion, the analysis of the SARS-CoV-2 introductions in Galicia suggests a disconnection between the number of introductions and the case numbers. The limited number of descendant cases from these introductions suggests that they did not markedly impact the evolution of the pandemic in Galicia. Furthermore, the source and frequency of these introductions appear to be influenced by the timing and nature of the restrictions implemented. Notably, coastal cities emerged as key nodes in the viral transmission networks, serving both as reservoirs for sustaining local transmission and as sources for the virus’s dispersion to smaller, more distant locations. This dynamic underscores the role of urban centers in the epidemiology of SARS-CoV-2, revealing how geographic, social, and mobility factors combine to shape the virus’s spread within and beyond these population hubs.

## Acknowledgments

We gratefully acknowledge all data contributors, i.e., the Authors and their Originating laboratories responsible for obtaining the specimens, and their Submitting laboratories for generating the genetic sequence and metadata and sharing via the GISAID Initiative, on which this research is based. This work was funded by grant EPICOVIGAL FONDO SUPERA-COVID19 from Banco Santander-CSIC-CRUE and grant CT850A-2 from ACIS SERGAS from the Consellería de Sanidade Xunta de Galicia. PGG was supported by grant ED481A-2021/345 from the Consellería de Cultura, Educación e Universidade Xunta de Galicia. SD acknowledges support from the *Fonds National de la Recherche Scientifique* (F.R.S.-FNRS, Belgium; grant no. F.4515.22). SD and GB acknowledge support from the Research Foundation - Flanders (*Fonds voor Wetenschappelijk Onderzoek* – *Vlaanderen*, FWO, Belgium; grant no. G098321N) and from the European Union Horizon RIA 2023 project LEAPS (grant no. 101094685). GB acknowledges support from the Internal Funds KU Leuven (Grant No. C14/18/094), from the Research Foundation - Flanders (*Fonds voor Wetenschappelijk Onderzoek* – *Vlaanderen*, FWO, Belgium; grant no. G0E1420N) and from the DURABLE EU4Health project 02/2023-01/2027, which is co-funded by the European Union (call EU4H-2021-PJ4; grant no. 101102733). SD and PL acknowledge support from the European Union Horizon 2020 project MOOD (grant agreement no. 874850). PL and MAS acknowledge support from the European Union’s Horizon 2020 research and innovation programme (grant agreement no. 725422 – ReservoirDOCS), from the Wellcome Trust through project 206298/Z/17/Z and from the National Institutes of Health grants R01 AI153044, R01 AI162611 and U19 AI135995. PL also acknowledges support from the Research Foundation – Flanders (*Fonds voor Wetenschappelijk Onderzoek - Vlaanderen*, G0D5117N, and G051322N); MIV, JCS and NSO acknowledge support from the Foundation for Science and Technology (FCT) (project UIDB/50026/2020, UIDP/50026/2020).

## Data availability

All genome sequences and associated metadata in this dataset are published in GISAID’s EpiCoV database. To view the contributors of each individual sequence with details such as accession number, Virus name, Collection date, Originating Lab, Submitting Lab, and the list of Authors, visit https://epicov.org/epi3/epi_set/240124ms (EPI_SET_ID: EPI_SET_240124ms, Supplementary Table 3).

## References

Aksamentov, Ivan, Cornelius Roemer, Emma B. Hodcroft, and Richard A. Neher. 2021. “Nextclade: Clade Assignment, Mutation Calling and Quality Control for Viral Genomes.” Journal of Open Source Software 6 (67): 3773.

Andrews, S. 2010. FastQC: A Quality Control Tool for High Throughput Sequence Data. http://www.bioinformatics.babraham.ac.uk/projects/fastqc/.

Argimón, Silvia, Khalil Abudahab, Richard J. E. Goater, Artemij Fedosejev, Jyothish Bhai, Corinna Glasner, Edward J. Feil, et al. 2016. “Microreact: Visualizing and Sharing Data for Genomic Epidemiology and Phylogeography.” Microbial Genomics 2 (11): e000093.

Ayres, Daniel L., Michael P. Cummings, Guy Baele, Aaron E. Darling, Paul O. Lewis, David L. Swofford, John P. Huelsenbeck, Philippe Lemey, Andrew Rambaut, and Marc A. Suchard. 2019. “BEAGLE 3: Improved Performance, Scaling, and Usability for a High-Performance Computing Library for Statistical Phylogenetics.” Systematic Biology 68 (6): 1052–61.

Bager, Peter, Jan Wohlfahrt, Samir Bhatt, Marc Stegger, Rebecca Legarth, Camilla Holten Møller, Robert Leo Skov, et al. 2022. “Risk of Hospitalisation Associated with Infection with SARS-CoV-2 Omicron Variant versus Delta Variant in Denmark: An Observational Cohort Study.” The Lancet Infectious Diseases 22 (7): 967–76.

Baniasad, Maryam, Morvarid Golrokh Mofrad, Bahare Bahmanabadi, and Sajad Jamshidi. 2021. “COVID-19 in Asia: Transmission Factors, Re-Opening Policies, and Vaccination Simulation.” Environmental Research 202 (111657): 111657.

Beca-Martínez, María Teresa, María Romay-Barja, María Falcón-Romero, Carmen Rodríguez-Blázquez, Agustín Benito-Llanes, and María João Forjaz. 2022. “Compliance with the Main Preventive Measures of COVID-19 in Spain: The Role of Knowledge, Attitudes, Practices, and Risk Perception.” Transboundary and Emerging Diseases 69 (4): e871–82.

Campbell, Finlay, Brett Archer, Henry Laurenson-Schafer, Yuka Jinnai, Franck Konings, Neale Batra, Boris Pavlin, et al. 2021. “Increased Transmissibility and Global Spread of SARS-CoV-2 Variants of Concern as at June 2021.” Eurosurveillance 26 (24): 2100509.

Carabelli, Alessandro M., Thomas P. Peacock, Lucy G. Thorne, William T. Harvey, Joseph Hughes, Sharon J. Peacock, Wendy S. Barclay, Thushan I. de Silva, Greg J. Towers, and David L. Robertson. 2023. “SARS-CoV-2 Variant Biology: Immune Escape, Transmission and Fitness.” Nature Reviews. Microbiology 21 (3): 162–77.

Carballada, Ángel Miramontes, and Jose Balsa-Barreiro. 2021. “Boletín de la Asociación Española de Geografía.” Boletín de la Asociación de Geógrafos Españoles, no. 91 (November). 10.21138/bage.3157.

Castelán-Sánchez, Hugo G., León P. Martínez-Castilla, Gustavo Sganzerla-Martínez, Jesús Torres-Flores, and Gamaliel López-Leal. 2022. “Genome Evolution and Early Introductions of the SARS-CoV-2 Omicron Variant in Mexico.” Virus Evolution 8 (2): veac109.

Dellicour, Simon, Samuel L. Hong, Bram Vrancken, Antoine Chaillon, Mandev S. Gill, Matthew T. Maurano, Sitharam Ramaswami, et al. 2021. “Dispersal Dynamics of SARS-CoV-2 Lineages during the First Epidemic Wave in New York City.” PLoS Pathogens 17 (5): e1009571.

Dellicour, Simon, Philippe Lemey, Marc A. Suchard, Marius Gilbert, and Guy Baele. 2022. “Accommodating Sampling Location Uncertainty in Continuous Phylogeography.” Virus Evolution 8 (1): veac041.

Dellicour, Simon, Rebecca Rose, Nuno R. Faria, Philippe Lemey, and Oliver G. Pybus. 2016. “SERAPHIM: Studying Environmental Rasters and Phylogenetically Informed Movements.” Bioinformatics 32 (20). 10.1093/bioinformatics/btw384.

Didier, Gustavo, Nathan E. Glatt-Holtz, Andrew J. Holbrook, Andrew F. Magee, and Marc A. Suchard. 2024. “On the Surprising Effectiveness of a Simple Matrix Exponential Derivative Approximation, with Application to Global SARS-CoV-2.” Proceedings of the National Academy of Sciences of the United States of America 121 (3): e2318989121.

Elbe, Stefan, and Gemma Buckland-Merrett. 2017. “Data, Disease and Diplomacy: GISAID’s Innovative Contribution to Global Health.” Global Challenges (Hoboken, NJ) 1 (1): 33–46.

Elliott, Paul, Barbara Bodinier, Oliver Eales, Haowei Wang, David Haw, Joshua Elliott, Matthew Whitaker, et al. 2022. “Rapid Increase in Omicron Infections in England during December 2021: REACT-1 Study.” Science 375 (6587): 1406–11.

Eurosurveillance editorial team. 2020. “Note from the Editors: World Health Organization Declares Novel Coronavirus (2019-nCoV) Sixth Public Health Emergency of International Concern.” Euro Surveillance: Bulletin Europeen Sur Les Maladies Transmissibles = European Communicable Disease Bulletin 25 (5). 10.2807/1560-7917.ES.2020.25.5.200131e.

Gill, Mandev S., Philippe Lemey, Shannon N. Bennett, Roman Biek, and Marc A. Suchard. 2016. “Understanding Past Population Dynamics: Bayesian Coalescent-Based Modeling with Covariates.” Systematic Biology 65 (6): 1041–56.

Gill, Mandev S., Philippe Lemey, Nuno R. Faria, Andrew Rambaut, Beth Shapiro, and Marc A. Suchard. 2013. “Improving Bayesian Population Dynamics Inference: A Coalescent-Based Model for Multiple Loci.” Molecular Biology and Evolution 30 (3): 713–24.

Grubaugh, Nathan D., Karthik Gangavarapu, Joshua Quick, Nathaniel L. Matteson, Jaqueline Goes De Jesus, Bradley J. Main, Amanda L. Tan, et al. 2019. “An Amplicon-Based Sequencing Framework for Accurately Measuring Intrahost Virus Diversity Using PrimalSeq and iVar.” Genome Biology 20 (1): 8.

Hadfield, James, Colin Megill, Sidney M. Bell, John Huddleston, Barney Potter, Charlton Callender, Pavel Sagulenko, Trevor Bedford, and Richard A. Neher. 2018. “Nextstrain: Real-Time Tracking of Pathogen Evolution.” Bioinformatics 34 (23): 4121–23.

Hasegawa, M., H. Kishino, and T. Yano. 1985. “Dating of the Human-Ape Splitting by a Molecular Clock of Mitochondrial DNA.” Journal of Molecular Evolution 22 (2): 160–74.

Hoang, Diep Thi, Olga Chernomor, Arndt von Haeseler, Bui Quang Minh, and Le Sy Vinh. 2018. “UFBoot2: Improving the Ultrafast Bootstrap Approximation.” Molecular Biology and Evolution 35 (2): 518–22.

Hodcroft, Emma B., Moira Zuber, Sarah Nadeau, Timothy G. Vaughan, Katharine H. D. Crawford, Christian L. Althaus, Martina L. Reichmuth, et al. 2021. “Spread of a SARS-CoV-2 Variant through Europe in the Summer of 2020.” Nature 595 (7869): 707–12.

Huska, Matthew R., and Denis Beslic. 2022. Breakfast - FAST outBREAK Detection and Sequence Clustering (version v0.4.3). https://github.com/rki-mf1/breakfast.

Ji, Xiang, Zhenyu Zhang, Andrew Holbrook, Akihiko Nishimura, Guy Baele, Andrew Rambaut, Philippe Lemey, and Marc A. Suchard. 2020. “Gradients Do Grow on Trees: A Linear-Time O(N)-Dimensional Gradient for Statistical Phylogenetics.” Molecular Biology and Evolution 37 (10): 3047–60.

Kaleta, Tamara, Lisa Kern, Samuel Leandro Hong, Martin Hölzer, Georg Kochs, Julius Beer, Daniel Schnepf, et al. 2022. “Antibody Escape and Global Spread of SARS-CoV-2 Lineage A.27.” Nature Communications 13 (1): 1152.

Kalyaanamoorthy, Subha, Bui Quang Minh, Thomas K. F. Wong, Arndt von Haeseler, and Lars S. Jermiin. 2017. “ModelFinder: Fast Model Selection for Accurate Phylogenetic Estimates.” Nature Methods 14 (6): 587–89.

Kraemer, Moritz U. G., Verity Hill, Christopher Ruis, Simon Dellicour, Sumali Bajaj, John T. McCrone, Guy Baele, et al. 2021. “Spatiotemporal Invasion Dynamics of SARS-CoV-2 Lineage B.1.1.7 Emergence.” Science 373 (6557): 889–95.

Lemey, Philippe, Nick Ruktanonchai, Samuel L. Hong, Vittoria Colizza, Chiara Poletto, Frederik Van den Broeck, Mandev S. Gill, et al. 2021. “Untangling Introductions and Persistence in COVID-19 Resurgence in Europe.” Nature 595 (7869): 713–17.

Li, Heng. 2013. “Aligning Sequence Reads, Clone Sequences and Assembly Contigs with BWA-MEM.” http://arxiv.org/abs/1303.3997.

Minh, Bui Quang, Heiko A. Schmidt, Olga Chernomor, Dominik Schrempf, Michael D. Woodhams, Arndt von Haeseler, and Robert Lanfear. 2020. “IQ-TREE 2: New Models and Efficient Methods for Phylogenetic Inference in the Genomic Era.” Molecular Biology and Evolution 37 (5): 1530–34.

Minin, Vladimir N., and Marc A. Suchard. 2008. “Counting Labeled Transitions in Continuous-Time Markov Models of Evolution.” Journal of Mathematical Biology 56 (3): 391–412.

O’Toole, Áine, Emily Scher, Anthony Underwood, Ben Jackson, Verity Hill, John T. McCrone, Rachel Colquhoun, et al. 2021. “Assignment of Epidemiological Lineages in an Emerging Pandemic Using the Pangolin Tool.” Virus Evolution 7 (2): veab064.

Paradis, Emmanuel, and Klaus Schliep. 2019. “Ape 5.0: An Environment for Modern Phylogenetics and Evolutionary Analyses in R.” Bioinformatics 35 (3): 526–28.

“Plan de Respuesta Temprana En Un Escenario de Control de La Pandemia Por COVID-19.” 2020. Ministerio de Sanidad. https://www.sanidad.gob.es/profesionales/saludPublica/ccayes/alertasActual/nCov/documentos/COVID19_Plan_de_respuesta_temprana_escenario_control.pdf.

Plessis, Louis du, John T. McCrone, Alexander E. Zarebski, Verity Hill, Christopher Ruis, Bernardo Gutierrez, Jayna Raghwani, et al. 2021. “Establishment and Lineage Dynamics of the SARS-CoV-2 Epidemic in the UK.” Science 371 (6530): 708–12.

Quick, Joshua, Nathan D. Grubaugh, Steven T. Pullan, Ingra M. Claro, Andrew D. Smith, Karthik Gangavarapu, Glenn Oliveira, et al. 2017. “Multiplex PCR Method for MinION and Illumina Sequencing of Zika and Other Virus Genomes Directly from Clinical Samples.” Nature Protocols 12 (6): 1261–76.

R Core Team. 2022. R: A Language and Environment for Statistical Computing. R Foundation for Statistical Computing, Vienna, Austria. https://www.R-project.org/.

Sagulenko, Pavel, Vadim Puller, and Richard A. Neher. 2018. “TreeTime: Maximum-Likelihood Phylodynamic Analysis.” Virus Evolution 4 (1): vex042.

Sharma, Mrinank, Sören Mindermann, Charlie Rogers-Smith, Gavin Leech, Benedict Snodin, Janvi Ahuja, Jonas B. Sandbrink, et al. 2021. “Understanding the Effectiveness of Government Interventions against the Resurgence of COVID-19 in Europe.” Nature Communications 12 (1): 1–13.

Shen, Wei, Shuai Le, Yan Li, and Fuquan Hu. 2016. “SeqKit: A Cross-Platform and Ultrafast Toolkit for FASTA/Q File Manipulation.” PloS One 11 (10): e0163962.

Suchard, Marc A., Philippe Lemey, Guy Baele, Daniel L. Ayres, Alexei J. Drummond, and Andrew Rambaut. 2018. “Bayesian Phylogenetic and Phylodynamic Data Integration Using BEAST 1.10.” Virus Evolution 4 (1): vey016.

Tegally, Houriiyah, Eduan Wilkinson, Joseph L-H Tsui, Monika Moir, Darren Martin, Anderson Fernandes Brito, Marta Giovanetti, et al. 2023. “Dispersal Patterns and Influence of Air Travel during the Global Expansion of SARS-CoV-2 Variants of Concern.” Cell 186 (15): 3277–90.e16.

Truong Nguyen, Phuoc, Ravi Kant, Frederik Van den Broeck, Maija T. Suvanto, Hussein Alburkat, Jenni Virtanen, Ella Ahvenainen, et al. 2022. “The Phylodynamics of SARS-CoV-2 during 2020 in Finland.” Communications Medicine 2 (1): 1–9.

Tsui, Joseph L-H, John T. McCrone, Ben Lambert, Sumali Bajaj, Rhys P. D. Inward, Paolo Bosetti, Rosario Evans Pena, et al. 2023. “Genomic Assessment of Invasion Dynamics of SARS-CoV-2 Omicron BA.1.” Science 381 (6655): 336–43.

Worldometer. 2020. “COVID-19 Virus Pandemic.” Worldometer. 2020. https://www.worldometers.info/coronavirus/.

Worobey, Michael, Joshua I. Levy, Lorena Malpica Serrano, Alexander Crits-Christoph, Jonathan E. Pekar, Stephen A. Goldstein, Angela L. Rasmussen, et al. 2022. “The Huanan Seafood Wholesale Market in Wuhan Was the Early Epicenter of the COVID-19 Pandemic.” Science 377 (6609): 951–59.

Yang, Z. 1994. “Maximum Likelihood Phylogenetic Estimation from DNA Sequences with Variable Rates over Sites: Approximate Methods.” Journal of Molecular Evolution 39 (3): 306–14.

