## Supplemental files for "Dispersal history of SARS-CoV-2 in Galicia, Spain"

#### Supplementary Material

**Table S1. List of kits used for extracting DNA/RNA from the nasopharyngeal exudates and measuring SARS-CoV-2 genome copy number through real-time RT-PCR.**

| |
|---|
| **Extraction kits** |
| GenoXtract® NA Extraction Kit (Hain Lifescience, Nehren, Germany) |
| MagCore® Viral Nucleic Acid Extraction Kit (Low PCR Inhibition) (RBC Bioscience, New Taipei City, Taiwan) |
| MagNA Pure 24 Total NA Isolation kit (Roche Diagnostics, Basel, Switzerland) |
| MagNA Pure Compact NA Isolation kit (Roche Diagnostics, Basel, Switzerland) |
| Maxwell® 16 Viral Total Purification Kit (Promega Corporation, Wisconsin, USA) |
| Maxwell® RSC Viral Total Nucleic Acid Purification Kit (Promega Corporation, Wisconsin, USA) |
| STARMag 96x4 Universal Cartridge Kit (Seegene Inc., Seoul, Republic of Korea) |
| STARMag 96x4 Viral DNA/RNA 200C Cartridge Kit (Seegene, Seoul, Republic of Korea) |
| TANBead® NA Extraction Kit (Taiwan Advanced Nanotech, Taiwan) |
| Virus DNA/RNA Extraction Kit (Biocomma, Guangdong, China) |
| **Real-time RT-PCR kits** |
| Alinity m Resp-4-Plex Assay (Abbott, Abbott Park, Illinois, USA) |
| Alinity m SARS-CoV-2 AMP Kit (Abbott, Abbott Park, Illinois, USA) |
| Allplex™ 2019-nCoV Assay (Seegene Inc., Seoul, Republic of Korea) |
| Allplex™ SARS-CoV-2 Assay (Seegene Inc., Seoul, Republic of Korea) |
| Allplex™ SARS-CoV-2 FluA/FluB/RSV Assay (Seegene, Seoul, Republic of Korea) |
| Allplex™ SARS-CoV-2 Master Assay (Seegene Inc., Seoul, Republic of Korea) |
| BIOFIRE® Respiratory 2.1 plus panel (bioMérieux, France) |
| Cobas® SARS-CoV-2 (Roche Diagnostics, Basel, Switzerland) |
| Cobas® SARS-CoV-2 & Influenza A/B (Roche Diagnostics, Basel, Switzerland) |
| Liaison® MDX Simplexa COVID-19 FluA/B Direct (DiaSorin, Saluggia, Italy) |
| LightMix® Sarbecovirus E-gene ModularDx (TIB Molbiol, Berlin, Germany) |
| LightMix® Sarbecovirus E-gene, S-gene, RpRd-gene, N-gene ModularDx (TIB Molbiol, Berlin, Germany) |
| SARS-CoV-2 RT PCR Kit (Vircell, Granada, Spain) |
| Simplexa® COVID-19 Direct (DiaSorin, Saluggia, Italy) |
| VIASURE SARS-CoV-2 Real Time PCR Detection Kit (CerTest Biotec, S.L., Zaragoza, Spain) |
| VIASURE SARS-CoV-2, Flu & RSV Real Time PCR Detection Kit (CerTest Biotec, S.L., Zaragoza, Spain) |
| Xpert® Xpress CoV-2/Flu/RSV plus (Cepheid Europe, France) |
| Xpert® Xpress SARS-CoV-2 (Cepheid Europe, France) |

**Table S2. List of ISO 3166-2 codes used in the circular migration flow plots and the complete name of the location.**

| Code ISO 3166-2 | Complete name |
| --- | --- |
| BR | Brazil |
| CH | Switzerland |
| CN | China |
| CO | Colombia |
| DK | Denmark |
| ES-AN | Andalusia |
| ES-AS | Asturias |
| ES-CB | Cantabria |
| ES-CM | Castile-La Mancha |
| ES-CN | Canary Islands |
| ES-CT | Catalonia |
| ES-PV | Basque Country |
| ES-VC | Valencian Community |
| IE | Ireland |
| LU | Luxembourg |
| NL | Netherlands |

**Table S3. GISAID data.**

### SUPPLEMENTAL TABLE

**Data Availability**

| | |
|---|---|
| GISAID Identifier: | EPI_SET_240124ms |
| doi: | 10.55876/gis8.240124ms |

All genome sequences and associated metadata in this dataset are published in GISAID's EpiCoV database. To view the contributors of each individual sequence with details such as accession number, Virus name, Collection date, Originating Lab and Submitting Lab and the list of Authors, visit 10.55876/gis8.240124ms

**Data Snapshot**

- EPI_SET_240124ms is composed of 86,142 individual genome sequences.
- The collection dates range from 2020-07-08 to 2022-06-02;
- Data were collected in 138 countries and territories;
- All sequences in this dataset are compared relative to hCoV-19/Wuhan/WIV04/2019 (WIV04), the official reference sequence employed by GISAID (EPI_ISL_402124). Learn more at https://gisaid.org/WIV04.

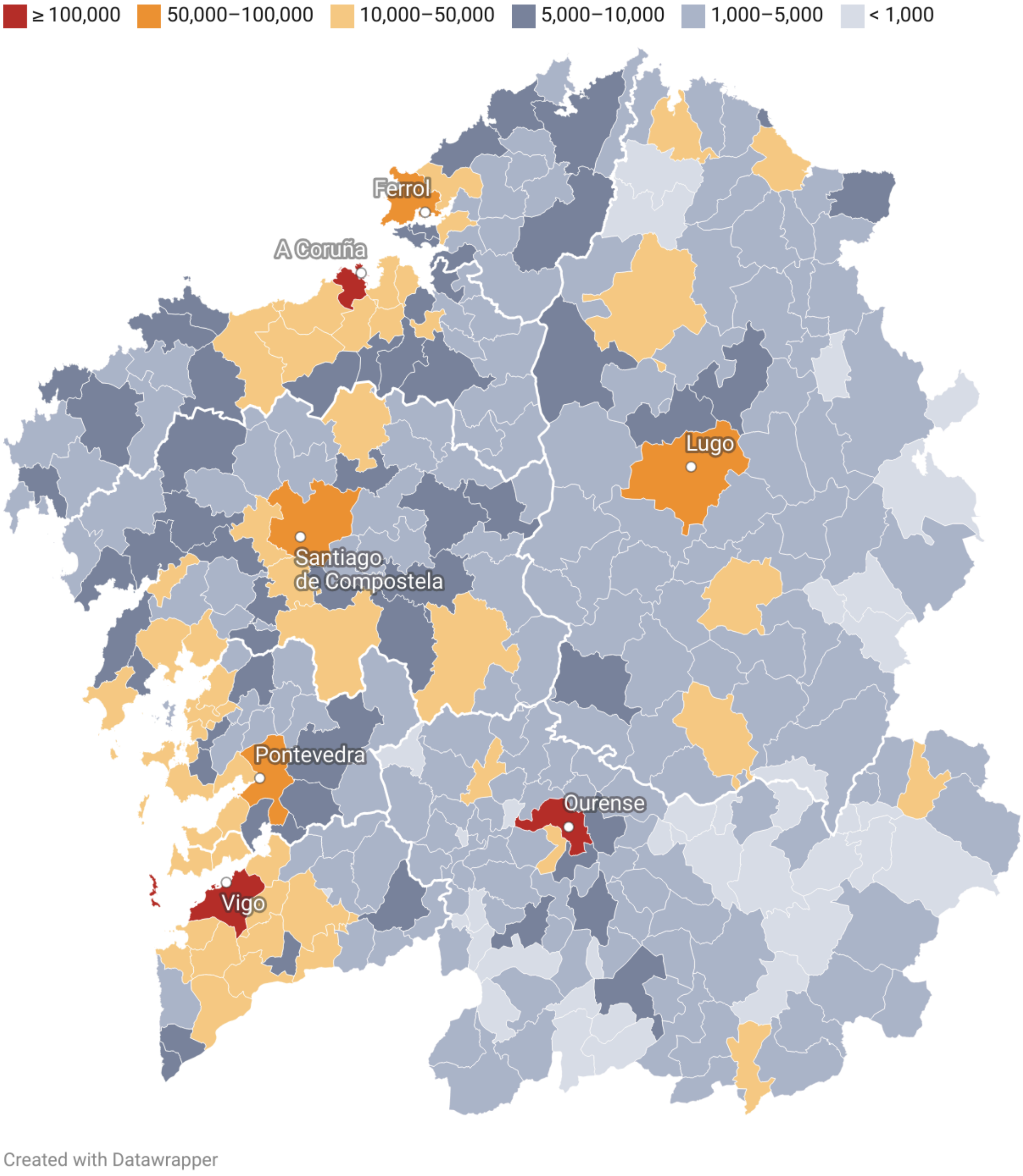


**Figure S1. Map of Galicia indicating the population of each municipality.** Thicker white lines delimit the seven healthcare areas. Data was taken from the Galician Statistics Institute (IGE).

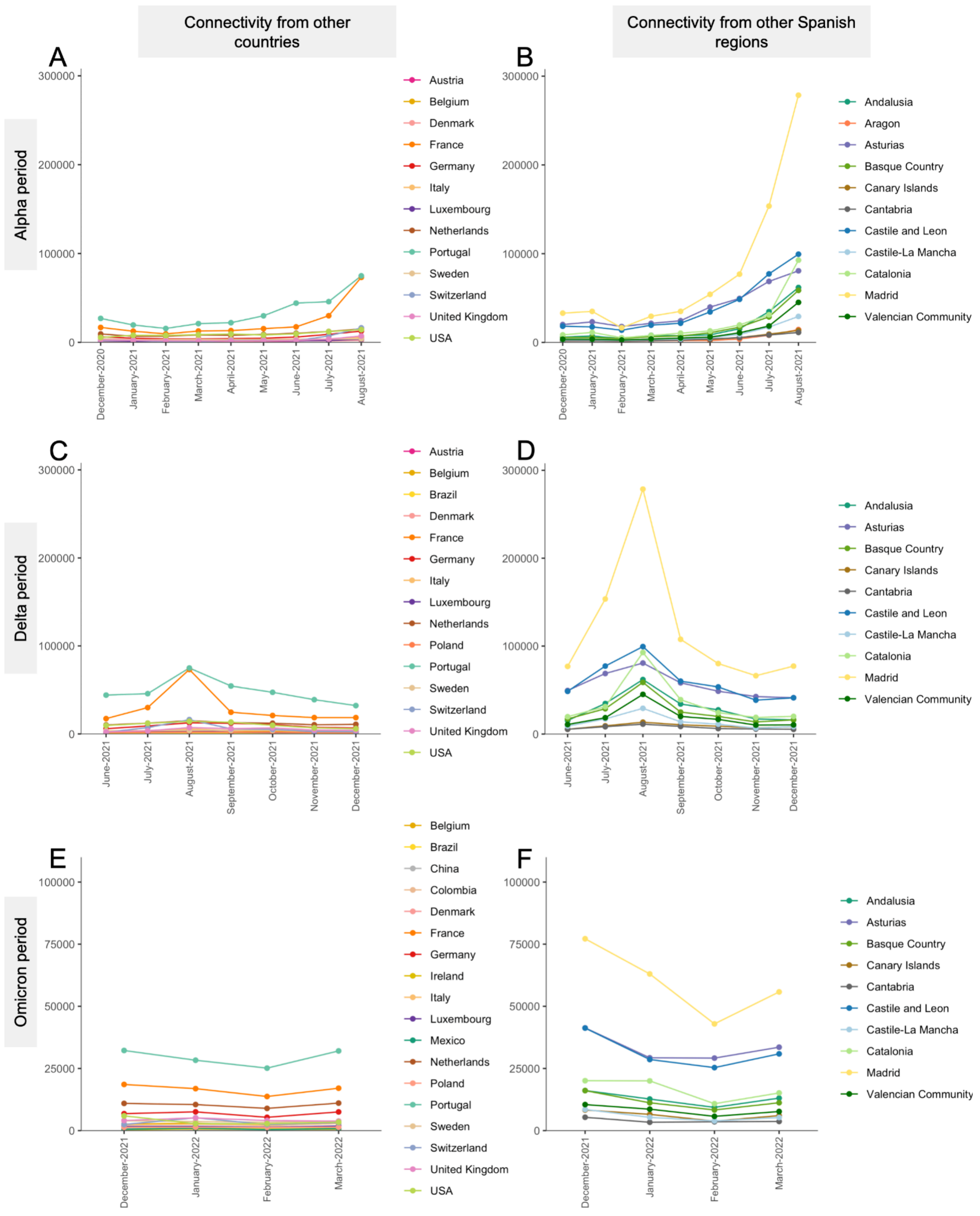


**Figure S2. Connectivity to Galicia.** Connectivity values were computed using the INE data for each period.

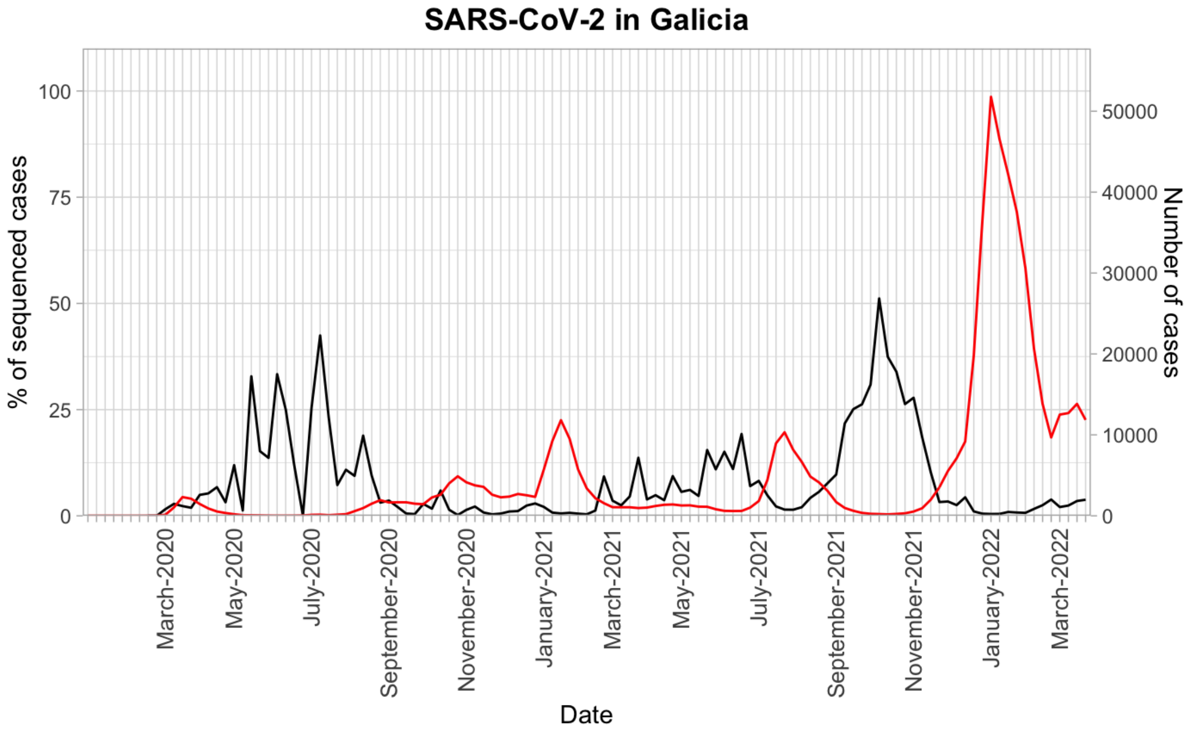


**Figure S3. Proportion of SARS-CoV-2 cases sequenced in Galicia along the epidemic waves from March 2020 to March 2022.** Proportion of cases sequenced (left-axis; black line) compared to the total number of cases reported in Galicia on the same date (right-axis; red line).

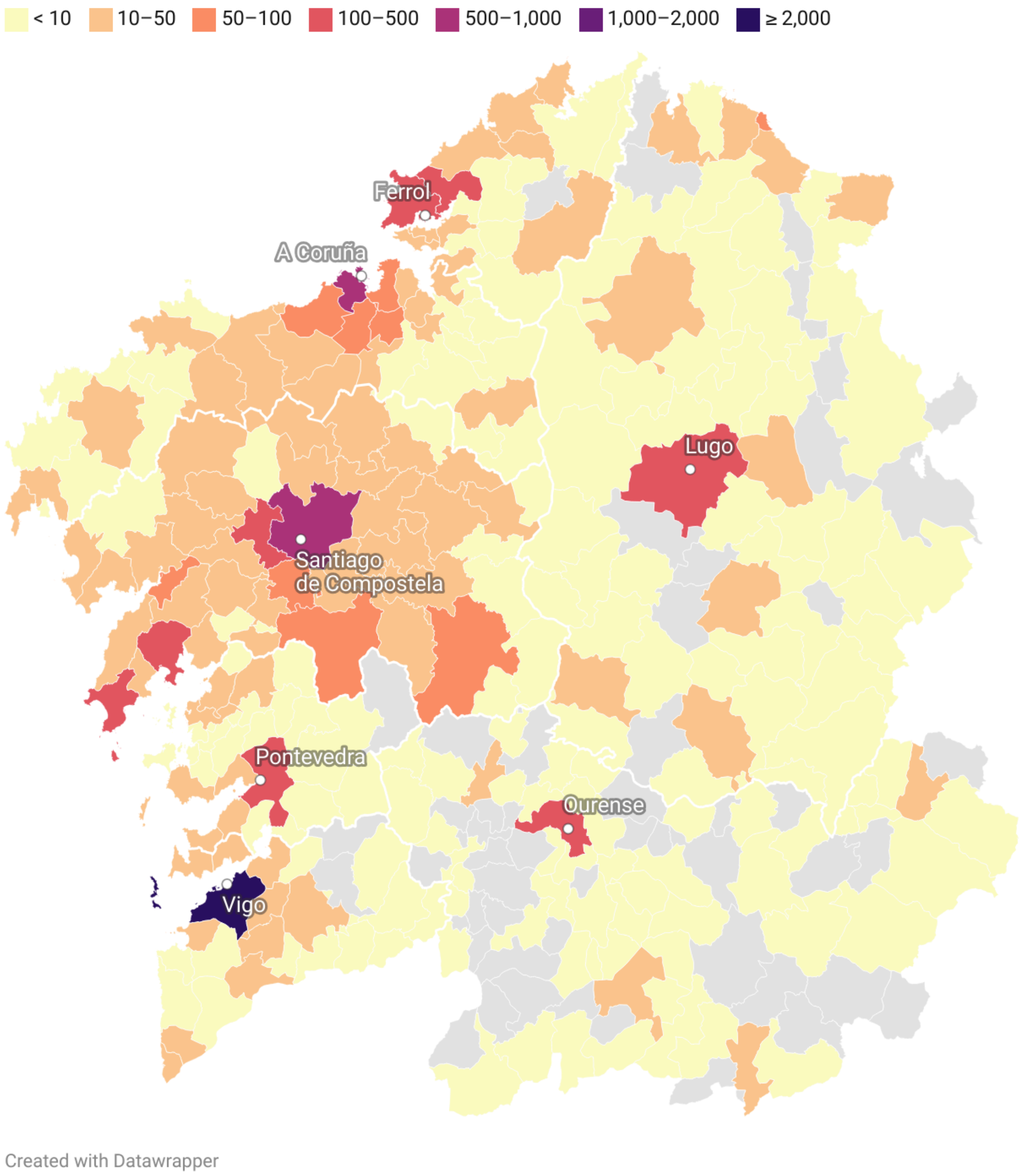


**Figure S4. Number of sequenced samples available per location in Galicia, March 2020 to March 2022.** The total number of samples includes those available in GISAID and those sequenced by the EPICOVIGAL Consortium.
